# Surviving Severe Acute Brain injury: Care trajectories and missed opportunities

**DOI:** 10.64898/2026.06.01.26354480

**Authors:** Adam L. Bunker, Ruth A. Engelberg, Robert G. Holloway, Claire J. Creutzfeldt

## Abstract

**INTRODUCTION:** Severe acute brain injury (stroke, traumatic brain injury or hypoxic-ischemic encephalopathy; SABI) is increasingly recognized as a chronic condition with care and communication needs beyond the initial hospitalization. This study aimed to characterize post-acute care patterns among SABI survivors, focusing on healthcare utilization and outpatient communication.

**METHODS:** Data were collected from a prospective cohort of hospitalized SABI patients using surveys, chart reviews, and the ED Information Exchange database. Socioeconomic disadvantage was assessed using the Area Deprivation Index (ADI), and qualitative analysis of outpatient notes examined conversations around palliative care needs and goals-of-care.

**RESULTS:** Two-thirds of patients (140/222) survived until discharge, primarily to nursing facilities (39%) or inpatient rehabilitation (38%). Among 109 with one-year follow-up, there were 89 hospitalizations, 104 ED visits, and 28 deaths. Patients from the most disadvantaged neighborhoods had significantly higher odds of rehospitalization or ED use within 30 days (OR 3.37, p=0.036). ADI was not linked to one-year utilization. Two-thirds of survivors (68%) were seen outpatient by primary care (40%), neurology/neurosurgery (57%), and palliative care (1%), but conversations rarely revisited prognosis or goals-of-care.

**CONCLUSIONS:** Our findings highlight the need for improved long-term care planning and communication, particularly for socioeconomically disadvantaged survivors of SABI.

## Introduction

Individuals with severe acute brain injury (SABI) who survive their hospitalization continue with high morbidity, mortality and healthcare utilization. Fragmented healthcare and frequent transitions in care create barriers to optimal communication regarding the patient’s treatment preferences or goals-of-care, increasing the risk for goal-discordant care, where treatment does not align with patient preferences.[1, 2] Risk factors for unplanned rehospitalization and Emergency Department (ED) utilization include age, prior stroke or TBI, and medical comorbidities, [3-6] but also non-medical drivers such as socioeconomic disadvantage.[7]

To build better models of post-acute care for SABI survivors, we need to better understand the frequency and predictors of rehospitalization or ED utilization, the receipt of care in outpatient settings following discharge, and the occurrence and content of patient-clinician conversations in the post-acute period.

The primary goal of this project was to describe the care trajectories for patients discharged alive from the hospital following SABI. The secondary goal was to explore outpatient clinician communication with patients and family regarding ongoing needs and treatment preferences during the year after injury. We hypothesized that survivors were more likely to return to ED or hospital if they came from areas of greater neighborhood disadvantage and if they were not seen in the outpatient setting in the post-acute period.

## Methods

Study participants had enrolled in the SuPPOrTT study, a single-center, prospective cohort study of patients with severe acute brain injury (SABI) admitted to the intensive care unit of a large academic hospital.[8] Eligibility criteria for the SuPPOrTT study included severe acute brain injury (SABI), defined as stroke (ischemic, intraparenchymal hemorrhage, or subarachnoid hemorrhage), traumatic brain injury, or hypoxic-ischemic encephalopathy (HIE) after cardiac arrest and a Glasgow Coma Scale score of ≤12 on or after hospital day 2. Patients were eligible for the SuPPOrTT study if they survived 48 hours or more and had at least one family member available to participate. The current analysis included patients from the SuPPOrTT cohort who survived to hospital discharge. Patients were dichotomized into traumatic (TBI) and vascular (stroke or HIE) SABI. As patients were unable to consent for themselves, consent was provided by their surrogate decision-maker on their behalf. The UW institutional review board approved the study.

### Data collection

We utilized prospectively collected data from the SuPPOrTT study, as well as additional retrospectively collected data from patient medical records, ED and hospitalization data from the Emergency Departments Information Exchange (EDIE)[9, 10] and block-group-level neighborhood disadvantage data from the Neighborhood Atlas Area Deprivation Index (ADI).[11, 12] EDIE is a health information exchange integrated into ED health records systems, centralizing data about patient visits to EDs and hospital admissions across states. Multiple states (Washington, Oregon, and Alaska) legislatively mandate that hospitals participate in such an information exchange, and all hospitals in these states utilize EDIE, allowing a richer dataset for readmissions beyond one hospital system.

### Outcome Measure

The primary outcome measure of interest was unplanned rehospitalization and ED utilization in the year following SABI. Because we assume multiple confounders of rehospitalization as time goes on, we examined early rehospitalization rates, which we defined as within 60 days after their SABI. Because the EDIE counts days after first admission rather than after discharge and the median length-of-stay for these patients was about one month, this “60 days after SABI” variable was closest to the well-established 30-day readmission rate.[13] Hospitalizations for planned procedures, e.g. cranioplasty, were not included.

### Predictors

Two predictors were of primary interest for factors associated with re-hospitalization and ED visits: (1) whether or not patients were seen by relevant providers in the outpatient setting and (2) patient’s neighborhood socioeconomic status at admission. Relevant providers included those in neurology, neurosurgery, physical medicine and rehabilitation (PM&R), primary care, palliative care. Visits were counted if they occurred between the date of discharge and the date of the patient’s first ED visit or rehospitalization; for those without any ED visits or rehospitalization, the end date was at one-year. Outpatient visits were dichotomized into any or no visits occurring during this period. For neighborhood socioeconomic status, we used the area deprivation index (ADI), a multidimensional evaluation a neighborhood’s education, employment, poverty, and housing quality from the Census Bureau’s American Community Survey. The patient’s address at the time of SABI was geocoded to the level of Census Block Group, with which state-specific ADI data were matched to patient data. ADI is expressed on a scale of 0-100 and split into quartiles for analysis, with the highest quartile indicating the highest level of deprivation.

### Statistical Analysis

To examine rehospitalizations and ED visits, patients were categorized into those with at least one unplanned rehospitalization or ED visit in the year after their SABI and those with neither unplanned rehospitalization nor ED visits during the same period. The same method was used to examine the period encompassing 60 days after their SABI. For comparisons, discrete variables were reported as counts and percentages and continuous variables as means and standard deviations. Proportions were compared using Pearson’s chi-square. Continuous variables were compared using two-sided student t-tests.

Multivariate logistic regression analyses to identify factors associated with unplanned rehospitalization and ED utilization included the following covariates regardless of statistical significance: age, gender, race, ethnicity, discharge disposition (home vs. other), and severity of brain injury (defined as GCS on hospital day 2). We included ED utilization and hospitalizations for the year prior to the index event as a way of controlling for other patterns of healthcare utilization. Outpatient visits and ADI quartiles were included as predictors of interest.

Analyses were conducted using Stata/BE 17.0 (StataCorp, College Park, TX, USA).

### Qualitative analysis

We extracted progress notes for all Neurology or Neurosurgery, primary care provider (PCP), PM&R or Palliative Care outpatient visits from the electronic health record (EHR). We specifically searched for any documentation of conversations around ongoing needs and treatment preferences. After a preliminary reading of documentation from the first 5 cases by the investigator team (AB, CJC), we devised a coding scheme to identify emergent themes and concepts addressed (or missing) in the clinic notes. The coding scheme was refined with subsequent cases through multiple readings by adding, removing, or grouping themes into primary thematic codes, each containing subcodes. We met monthly to discuss cases and refine the coding scheme.

### Data Availability

Anonymized data from this study are available upon request.

## Results

The SuPPOrTT study enrolled 222 patients, of whom 82 died in-hospital. The one-year trajectory for the 140 survivors is presented as an alluvial diagram in Figure 1. Thirty-one hospital survivors opted out of further data collection (additional chart review and use of EDIE) leaving 109 SABI survivors for this analysis. Overall, patients for whom the full 1-year data was available had a mean age of 58.6 years, 74.3% were white, and 17 patients (15.6%) came from neighborhoods with the highest ADI (see table 1). Most survivors (86.2%) had not been hospitalized within this system prior to the index hospitalization. Average GCS at enrollment was 8.0 (SD=2.6); 64.2% (n=70) had suffered vascular SABI (stroke or HIE). Patients for whom additional chart review and use of EDIE data were not available (n=31) were disproportionately non-white (35.4% vs 25.7% for those with additional data), Hispanic (12.9% vs 6.4%), and male (67.7% vs. 53.2%), but none of these differences were statistically significant.

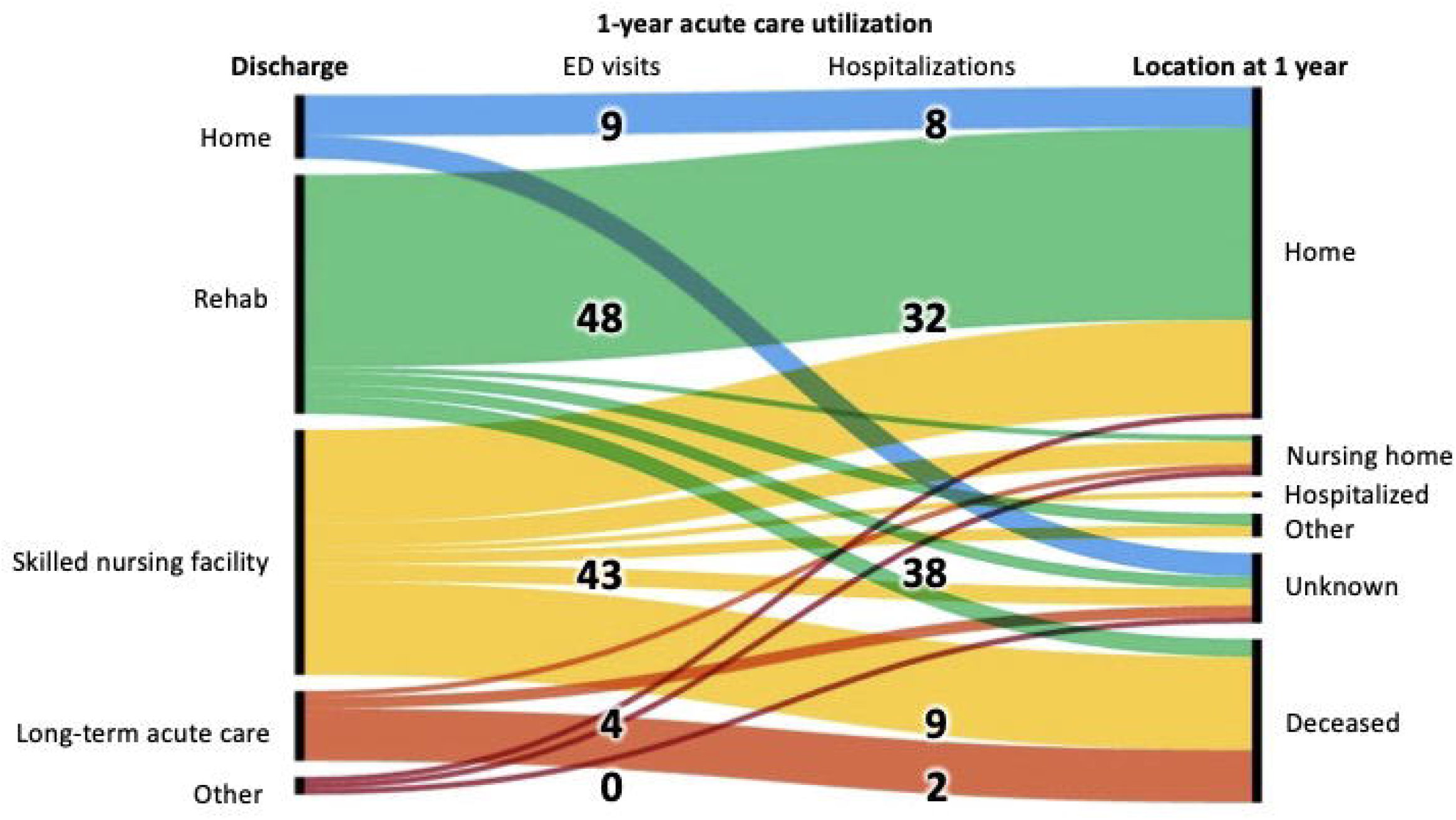

**Table 1:**
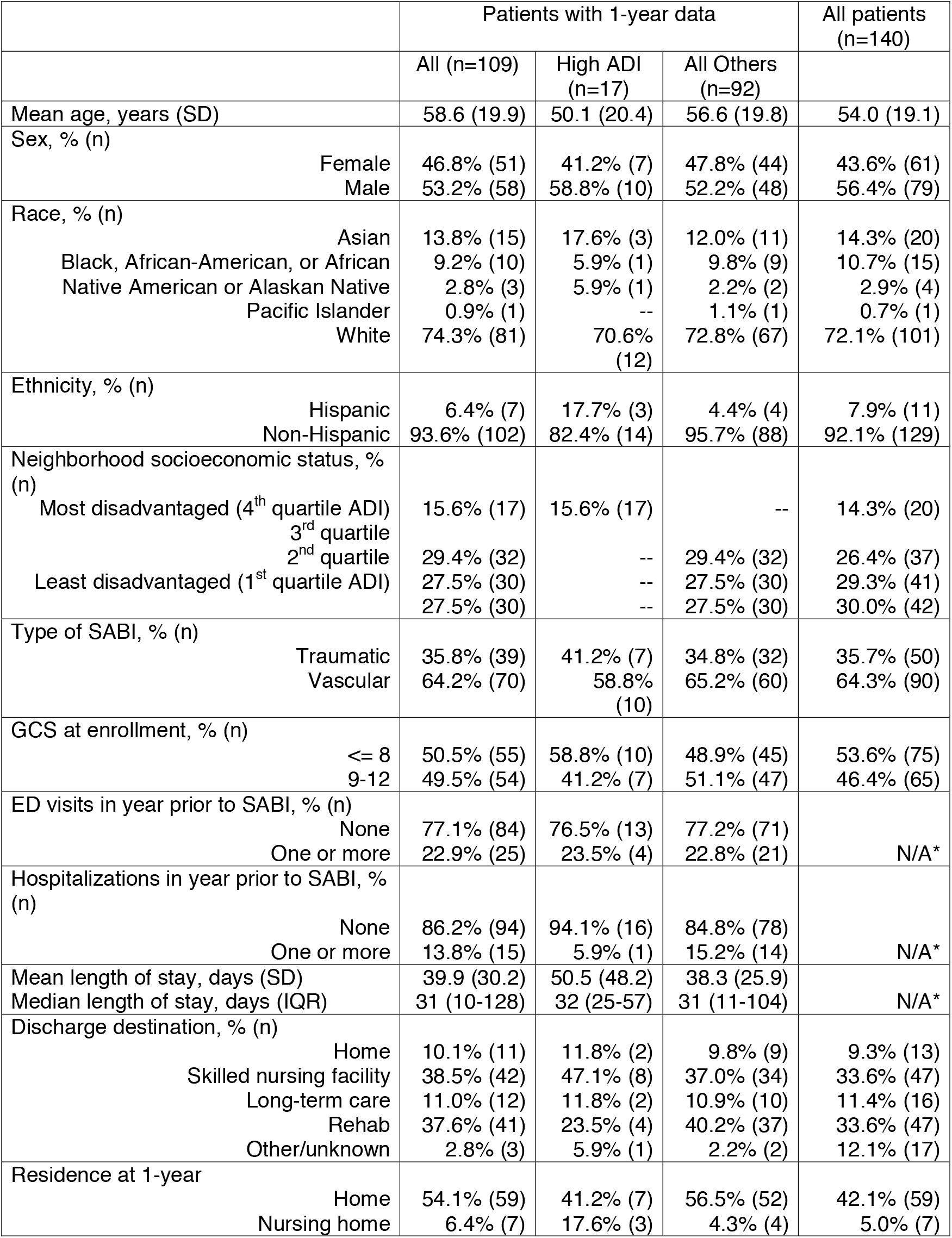
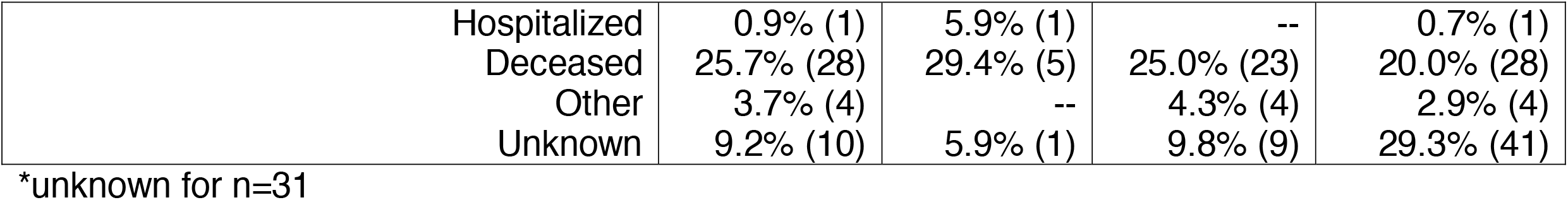
Patient Characteristics.

|  |  | Patients with 1-year data |  |  | All patients<br>(n=140) |
| --- | --- | --- | --- | --- | --- |
|  |  | All (n=109) | High ADI<br>(n=17) | All Others<br>(n=92) |  |
| Mean age, years (SD) |  | 58.6 (19.9) | 50.1 (20.4) | 56.6 (19.8) | 54.0 (19.1) |
| Sex, % (n) | Female | 46.8% (51) | 41.2% (7) | 47.8% (44) | 43.6% (61) |
|  | Male | 53.2% (58) | 58.8% (10) | 52.2% (48) | 56.4% (79) |
| Race, % (n) | Asian | 13.8% (15) | 17.6% (3) | 12.0% (11) | 14.3% (20) |
|  | Black, African-American, or African | 9.2% (10) | 5.9% (1) | 9.8% (9) | 10.7% (15) |
|  | Native American or Alaskan Native | 2.8% (3) | 5.9% (1) | 2.2% (2) | 2.9% (4) |
|  | Pacific Islander | 0.9% (1) | -- | 1.1% (1) | 0.7% (1) |
|  | White | 74.3% (81) | 70.6%<br>(12) | 72.8% (67) | 72.1% (101) |
| Ethnicity, % (n) | Hispanic | 6.4% (7) | 17.7% (3) | 4.4% (4) | 7.9% (11) |
|  | Non-Hispanic | 93.6% (102) | 82.4% (14) | 95.7% (88) | 92.1% (129) |
| Neighborhood socioeconomic status, %<br>(n) |  |  |  |  |  |
| Most disadvantaged (4<sup>th</sup> quartile ADI) |  | 15.6% (17) | 15.6% (17) | -- | 14.3% (20) |
| 3<sup>rd</sup> quartile |  |  |  |  |  |
| 2<sup>nd</sup> quartile |  | 29.4% (32) | -- | 29.4% (32) | 26.4% (37) |
| Least disadvantaged (1<sup>st</sup> quartile ADI) |  | 27.5% (30) | -- | 27.5% (30) | 29.3% (41) |
|  |  | 27.5% (30) | -- | 27.5% (30) | 30.0% (42) |
| Type of SABI, % (n) | Traumatic | 35.8% (39) | 41.2% (7) | 34.8% (32) | 35.7% (50) |
|  | Vascular | 64.2% (70) | 58.8%<br>(10) | 65.2% (60) | 64.3% (90) |
| GCS at enrollment, % (n) | <= 8 | 50.5% (55) | 58.8% (10) | 48.9% (45) | 53.6% (75) |
|  | 9-12 | 49.5% (54) | 41.2% (7) | 51.1% (47) | 46.4% (65) |
| ED visits in year prior to SABI, % (n) |  |  |  |  |  |
| None |  | 77.1% (84) | 76.5% (13) | 77.2% (71) | N/A* |
| One or more |  | 22.9% (25) | 23.5% (4) | 22.8% (21) |  |
| Hospitalizations in year prior to SABI, %<br>(n) |  |  |  |  |  |
| None |  | 86.2% (94) | 94.1% (16) | 84.8% (78) | N/A* |
| One or more |  | 13.8% (15) | 5.9% (1) | 15.2% (14) |  |
| Mean length of stay, days (SD) |  | 39.9 (30.2) | 50.5 (48.2) | 38.3 (25.9) | N/A* |
| Median length of stay, days (IQR) |  | 31 (10-128) | 32 (25-57) | 31 (11-104) |  |
| Discharge destination, % (n) |  |  |  |  |  |
| Home |  | 10.1% (11) | 11.8% (2) | 9.8% (9) | 9.3% (13) |
| Skilled nursing facility |  | 38.5% (42) | 47.1% (8) | 37.0% (34) | 33.6% (47) |
| Long-term care |  | 11.0% (12) | 11.8% (2) | 10.9% (10) | 11.4% (16) |
| Rehab |  | 37.6% (41) | 23.5% (4) | 40.2% (37) | 33.6% (47) |
| Other/unknown |  | 2.8% (3) | 5.9% (1) | 2.2% (2) | 12.1% (17) |
| Residence at 1-year | Home | 54.1% (59) | 41.2% (7) | 56.5% (52) | 42.1% (59) |
|  | Nursing home | 6.4% (7) | 17.6% (3) | 4.3% (4) | 5.0% (7) |

|  |  |  |  |  |
| --- | --- | --- | --- | --- |
| Hospitalized | 0.9% (1) | 5.9% (1) | -- | 0.7% (1) |
| Deceased | 25.7% (28) | 29.4% (5) | 25.0% (23) | 20.0% (28) |
| Other | 3.7% (4) | -- | 4.3% (4) | 2.9% (4) |
| Unknown | 9.2% (10) | 5.9% (1) | 9.8% (9) | 29.3% (41) |

\*unknown for n=31

### Care trajectories

Most patients were discharged to either a skilled nursing facility (38.5%) or inpatient rehabilitation (37.6%). Only 10.1% discharged home, with the remainder (11.0%) discharging to long-term care or other facilities. Within one year, one-quarter of patients had died (n=28, 25.7%), 10 of those (35.7%) in a hospital setting.

In the year following the initial hospitalization, 89 unplanned hospitalizations and 104 standalone ED visits occurred at hospitals within the EDIE system (Table 2). Almost one-half of survivors (n=48, 44%) had at least one unplanned hospitalization, and 47.7% (n=52) had at least one standalone ED visit. Over one-half (n=62, 56.9%) had a specialist follow-up visit with Neurology or Neurosurgery, and 44 (40.4%) saw a primary care provider within our system (Table 2). Among the 74 patients seen in the outpatient setting, 25 were able to communicate independently, all others had cognitive or communication impairment ranging from word finding difficulties to unable to communicate at all (Supplemental Table 1). One-third (n=35, 32.1%) did not have a clinic visit by a relevant provider in our system.

**Table 2:** Healthcare utilization after SABI among 109 survivors.

|  | **Total Visits, n**<br>**(%)** | **Unique Patients, n**<br>**(% of total 109**<br>**patients)** |
| --- | --- | --- |
| Outpatient visits* (1 year) | 528 (ref) | 74 (67.9%) |
| Neurology / Neurosurgery | 158 (29.9%) | 62 (56.9%) |
| Primary Care | 237 (44.9%) | 44 (40.4%) |
| Rehabilitative Medicine | 132 (25.0%) | 39 (35.8%) |
| Palliative Care | 1 (0.2%) | 1 (0.9%) |
| Total unplanned hospitalizations |  |  |
| Within 1 year | 89 (ref) | 48 (44.0%) |
| Within 60 days | 13 (14.6%) | 10 (9.2%) |
| Total standalone ED visits |  |  |
| Within 1 year | 104 (ref) | 52 (47.7%) |
| Within 60 days | 15 (14.4%) | 11 (10.1%) |

\*Outpatient visits that we considered relevant to SABI, including Neurology for Stroke or cardiac arrest, Neurosurgery for TBI, Primary care, Rehab Medicine and Palliative care.

Among those experiencing a return to hospital or ED (n=69), mean time between admission and rehospitalization was 65 days (SD=88) overall. For patients with any outpatient visits within the year following admission (n=54), mean time between admission and rehospitalization was 77 days (95% CI: 49.9-105.0) vs. 19 days (95% CI: 5.9-32.0) for those who were never seen (n=15) (two-sample t test=-2.25, p=0.028).

### Factors associated with early rehospitalization and ED use

After adjusting for age, gender, race, ethnicity, and prior ED/hospital utilization in multivariate analysis, patients living in socioeconomically disadvantaged neighborhoods had more than 3 times the odds of rehospitalization or ED use in the 60 days following admission (OR 3.37, 95% CI 1.08-10.52, p=0.036) compared to those living elsewhere. There were no differences in odds of early return to hospital or ED based on receipt of outpatient care (Table 3). Comparing odds of rehospitalization or ED use within one year identified no statistically significant differences across any groups.

**Table 3:** Patient characteristics and odds of early rehospitalization or ED utilization within 60 days from SABI admission.

|  | Univariate |  | Multivariate |  |
| --- | --- | --- | --- | --- |
|  | OR (95% CI) | p-value | OR (95% CI) | p-value |
| Age, over 60 | 1.26 (0.59-2.71) | 0.551 | 1.62 (0.70-3.77) | 0.261 |
| Gender, female | 0.99 (0.46-2.13) | 0.983 | 1.14 (0.49-2.65) | 0.753 |
| Race, non-white | 1.09 (0.46-2.60) | 0.845 | 0.97 (0.37-2.53) | 0.946 |
| Ethnicity, Hispanic | 1.98 (0.42-9.33) | 0.386 | 1.38 (0.25-7.72) | 0.717 |
| GCS score $\geq 9$ at enrollment | 0.82 (0.38-1.76) | 0.615 | 0.86 (0.36-2.01) | 0.72 |
| Hospitalization or ED utilization in prior year | 1.07 (0.47-2.45) | 0.873 | 1.21 (0.50-2.91) | 0.669 |
| Discharged to home | 0.50 (0.13-2.00) | 0.327 | 0.43 (0.09-2.01) | 0.286 |
| At least one clinic visit before hospitalization | 1.29 (0.56-2.94) | 0.546 | 1.62 (0.66-4.01) | 0.296 |
| **Disadvantaged socioeconomic environment (highest quartile ADI)** | **3.13 (1.06-9.22)** | **0.039** | **3.60 (1.13-11.44)** | **0.030** |

### Qualitative analysis

Three areas of missed opportunity emerged from our analysis of outpatient visit notes in the year after SABI. The first missed opportunity was deeper exploration of patient and family member well-being. The second was an unmet need for goals-of-care conversations. Third, patients and families’ questions about updated prognosis went mostly unanswered.

#### Patient and family member well-being

Patient well-being was frequently addressed in terms of the patient’s functional status including activities of daily living (ADLs) and level of dependence (“remains with expressive aphasia and R face, arm, leg weakness… mRS 4”). In the rare cases that the patient’s emotional well-being or mental health was discussed, family members reported substantial suffering:

> *“Husband reports that she has ‘dark days*.*’ States that life is not worth living, wished she had died in the accident, feels that she is a burden to her family. Denies thoughts of self-harm. Her husband is asking if she can see a mental health counselor*.*”* (Primary Care Provider)
>
> *“Family is not sure if [patient] is more irritable lately or just that she’s more interactive and frustrated by her physical limitations*.*”* (Neurology Provider)
>
> *“[Patient’s mother] states that just yesterday, [patient] ‘had a breakdown,’ was tearful, and questioned why he was still alive*.*”* (Rehabilitative Medicine Provider)

Comments like those above as well as positive depression screening using the patient health questionnaire did not reliably translate to further assessment or plan.

> *“Overall she seems depressed but given her severe impairments, some adjustment would be expected*.*”* (Primary Care Provider)

Assessment of family member well-being was similarly infrequent and focused on the practical aspects of caregiving.

> *“Placed referral to social work* .. *she should get a call to talk over caregiver hiring*⍰*and AFH options*.*”* (Rehabilitative Medicine Provider)
>
> *“…reports that things are going fairly well at home but he is really the primary caregiver*… *trying to apply for [disability]*.*”* (Rehabilitative Medicine Provider)

#### Patient-centered values and goals-of-care

Goals-of-care conversations occurred when a specific treatment was imminent.

> *“I talked frankly with her husband and daughter about goals of care*.. *I don’t think [draining the subdural] would make her any better… with horrible stroke, aphasia, hemiparesis, and poor quality-of-life they might wish to change goals of care*.*”* (Neurosurgery Provider)
>
> *“I know his function is low and yet family is pushing hard to have the plasty although it carries some risks and won’t make the patient better*.*”* (Neurosurgery Provider)

Among all 527 notes, only two revealed evidence of broader conversations to understand and revisit patient and family values:

> *“[Patient’s mother] knows that [patient] would not do well if she became very sick, and so does not see a point in overly heroic measures such as intubation or cardiac resuscitation.* ⍰*She would want [patient] to be hospitalized, though, should she suffer a pneumonia or other and receive antibiotics and other necessary treatment*.*”* (Neurology Provider)
>
> *“We also talked about*⍰*his… ICU/hospital stay, about the decisions that were made and about the care that is important to him going forward…*.*He actually thinks that he is better than he had expected to be and is optimistic about his progress. He… would still want aggressive measures should something unexpected happen. When I asked him whether he would make the same decisions again, he responded with ‘I don’t know*.*’”* (Neurology Provider)

#### Need for Updated Prognosis

Reassessment of prognosis or conversations about what to expect going forward were often desired by family members and evidently challenging for clinicians.

> *“Pt’s wife also had a number of questions about … the anticipated trajectory of recovery*. ⍰*We briefly reviewed his initial CT scan…however, I deferred most of the questions to the rehab visit they have tomorrow*.*”* (Neurology Provider, 6 months after hospitalization; of note, the Rehab visit that following day did not address the potential for recovery or anticipated trajectory)
>
> *“…began to ask questions about the patient’s prognosis and recovery potential.* ⍰*Some of these questions included whether or not this patient would walk again, to communicate again and what were realistic expectations for the patient… I was not prepared to delve into these conversations in detail but did explain that he suffered a major brain injury and he may have some slight improvements, but will likely always have a permanent neurologic disability and will not be the way he was before his injury*…*continued to ask questions specifically surrounding issues of long-term decision making. I apologized I was not prepared to discuss this*.*”* (Neurosurgery Provider 11 months after hospitalization)
>
> *“Patient has had long and bumpy road after his brain injury…. It is still very difficult to prognosticate what his long term physical and cognitive function will look like as he has not at any point stabilized for any significant length of time*.*”* (Rehabilitative Medicine Provider at 6 months after hospitalization)

From notes, it was sometimes unclear whether the information was shared with the family or not:

> *“Chance of functional recovery is poor at this point with no significant recovery seen since accident*.*”* (Neurology Provider)

## Discussion

This analysis of survivors of SABI suggests continued challenges for patients and their families that are insufficienly addressed in the post-acute period. Rehospitalizations were common, especially among those from socio-economically disadvantaged neighborhoods. Despite a high level of morbidity and mortality, goals-of-care conversations were rare in the outpatient setting.

Our findings are consistent with prior literature suggesting an association between socioeconomic status and re-admission. Re-admission diagnoses are unknown, but this finding clearly calls for a higher need for support among those already disadvantaged. Previous studies of patients with stroke have shown associations between readmission and the patient’s initial discharge destination. The lack of association with discharge destination in our study may be due to selection bias, as our cohort included only patients with severe acute brain injury and a relatively small proportion of home discharges (10% discharged home in our cohort vs. over one half in other studies[3, 14]). Previous research has found disparities in withdrawal of life-sustaining treatment across racial, ethnic, and neighborhood socioeconomic status groups.[15] One potential explanation for our finding greater odds of readmission among those who are most disadvantaged may relate to this prior research: if patients were less likely to have withdrawal of life-sustaining treatment, they would then be discharging with greater disease burden and needs, which may then manifest as increased odds of readmission.

Qualitative analysis revealed several opportunities to improve post-acute care for SABI survivors. We suggest that outpatient visits should routinely include (1) assessment of both patient and family emotional well-being; (2) re-visiting goals-of-care; and (3) conversations about the future. In the acute setting after SABI, prognostic uncertainty is a well-known and widely studied challenge, especially when families are tasked to make value-laden treatment decisions.[16-18] Life-sustaining treatments are often instituted during the acute hospitalization as a time-limited trial,[19] to allow additional time for prognosis to become clearer.[20] However, our study suggests that these decisions were rarely revisited. In the post-acute setting, most treatment decisions may not be imminent but patients and families still want to know what to expect. Prognostic uncertainty does decrease over time, and the degree of expected recovery is likely to become evident. In our analysis, when patients and families sought updated prognoses, it was not uncommon for their requests to be deferred to a future visit or different provider. Whether or not patients and families are in the spot they had either hoped or expected in the year after injury, their values, preferences, and ultimately, goals-of-care may evolve as they are now living with their new reality.[21] Considering the resources spent in the acute care of patients with SABI, there is an urgent need for healthcare systems to invest in ongoing post-acute care and support for these patients and their families. Psychosocial and practical support as well as re-evaluation of prognosis and treatment preferences may improve patient and family quality-of-life, reduce unwanted healthcare utilization and move us closer in our quest to provide patient-centered, goal-concordant care.

### Limitations

Several limitations should be considered. Our method of retrospective review of chart records to identify the content of post-SABI follow-up is inherently limited to include only those discussions that were documented by the provider. Even when providers self-report having had a goals of care conversations, more than 1 in 3 of these discussions are unidentifiable by only EHR documentation.[22] Additionally, it is possible that some patients went to providers who did not provide EHR data to our sources. This likely resulted in an underestimate of the number of patients who were seen by providers in the year after SABI. The magnitude of this underestimate was likely greatest for primary care as there are more primary care providers unaffiliated with large systems than, for example, neurosurgeons or neurologists. Finally, the single-center nature of this study and small sample size may limit generalizability to other settings.

## Data Availability

All data produced in the present study are available upon reasonable request to the authors

## Supplementary Tables

**Table S1:** Patient ability to Communicate (with chart note examples)

|  | ***n*** | ***Examples*** |
| --- | --- | --- |
| *Unable to communicate at all* | 7 |  |
| *Yes / No only* | 11 | *"He continues to be mute with occasional small vocalizations, □ but is able to communicate with movements of his eyes, □ and appears to understand emotions, such as when his father-in-law recently passed away and he was noted to cry." (neuro)* |
|  |  | *"He is cognizant but can not verbalize, able to answer questions with nod of head and right hand gestures." (PCP)* |
|  |  | *"Patient is aphasic but nods yes/no to most questions. □ Per his caregiver, he understands what people are saying and gives appropriate yes/no answers." (cardiology)* |
|  |  | *"she seems to be doing well and is able to communicate by moving her eyes vertically" (PCP)* |
|  |  | *"The patient is able to express herself through yes and no responses, and states that there is problem with her eyesight, but is unable to elaborate." (neurosurg)* |
| *Communicates beyond yes/no, but with significant difficulty* | 14 | *"non-fluent aphasia and exceedingly poor memory"* |
|  |  | *"Answers very simple questions, but language is garbled and difficult to understand, oriented to self only" (neurosurg)* |
| *Communicates with some difficulty (e.g. mild cognitive impairment or aphasia)* | 17 | *"[has] episodes of knowing what he would like to say, however not being able to get the correct words out."* |
|  |  | *"residual right sided weakness and communication impairment which have been improving since his return home."* |
| *Able to fully communicate* | 25 |  |

**Table S2:** Patient characteristics and odds of rehospitalization or ED utilization within 1 year.

|  | **Univariate** |  | **Multivariate** |  |
| --- | --- | --- | --- | --- |
|  | **OR (95% CI)** | **p-value** | **OR (95% CI)** | **p-value** |
| **Age, over 60** | 0.79 (0.37-1.69) | 0.541 | 0.82 (0.36-1.83) | 0.625 |
| **Gender, female** | 0.86 (0.40-1.84) | 0.696 | 0.81 (0.36-1.82) | 0.610 |
| **Race, non-white** | 1.15 (0.62-3.68) | 0.360 | 1.54 (0.59-3.98) | 0.375 |
| **Ethnicity, Hispanic** | 1.01 (0.22-4.75) | 0.988 | 0.95 (0.17-5.24) | 0.954 |
| **GCS score <sup>3</sup> 9 at enrollment** | 1.04 (0.49-2.23) | 0.912 | 1.23 (0.54-2.82) | 0.627 |
| **Disadvantaged socioeconomic environment** | 2.02 (0.66-6.19) | 0.220 | 1.91 (0.58-6.27) | 0.286 |
| **Hospitalization or ED utilization in prior year** | 1.25 (0.54-2.86) | 0.605 | 1.21 (0.52-2.85) | 0.660 |
| **Discharged to home** | 0.90 (0.26-3.15) | 0.869 | 0.93 (0.23-3.71) | 0.918 |
| **At least one clinic visit before hospitalization** | 0.86 (0.40-1.84) | 0.701 | 0.86 (0.37-1.99) | 0.731 |

## Notes

Statements and Declarations: This work was supported by a grant from the National Institute of Neurological Disorders and Stroke (Award No. K23NS099421).

### Competing Interest Statement

This work was supported by a grant from the National Institute of Neurological Disorders and Stroke (Award No. K23NS09

### Author Declarations

The University of Washington institutional review board approved the study.

